# PREVALENCE OF TUBERCULOSIS IN CENTRAL ASIA AND SOUTHERN CAUCASUS: A SYSTEMATIC LITERATURE REVIEW

**DOI:** 10.1101/2025.04.28.25326554

**Authors:** Malika Idayat, Nailya Kozhekenova, Oyunzul Amartsengel, Kamila Akhmetova, Elena Von Der Lippe, Natalya Glushkova

## Abstract

In 2023, tuberculosis (TB) caused an estimated 1.25 million deaths globally, including 161,000 among people with HIV. After being temporarily surpassed by COVID-19, TB has likely returned as the leading cause of death from a single infectious agent. Central Asia and the Southern Caucasus remain high-burden regions, with Mongolia showing the highest TB prevalence. This systematic review aimed to synthesize current data on TB prevalence in Central Asia, Southern Caucasus, and Mongolia to support public health strategies and research priorities. A comprehensive search of PubMed and Google Scholar was conducted for English-language articles published up to 2023. Studies were assessed using a modified Newcastle-Ottawa Scale. Nine studies met inclusion criteria, covering Kazakhstan, Kyrgyzstan, Uzbekistan, Tajikistan, Turkmenistan, Mongolia, Georgia, Armenia, and Azerbaijan. TB incidence ranged from 67 per 100,000 in Kazakhstan to 190 per 100,000 in Kyrgyzstan, with a prevalence of 68.5% in Mongolia. TB affected men more frequently (65.3%), with key risk factors including HIV (30.5%), comorbidities, and undernutrition. Drug-resistant TB imposed a significant economic burden, with treatment costs ranging from $106 to $3,125. Strengthening surveillance, improving data collection, and conducting longitudinal studies are essential for designing effective TB control strategies in these regions.

## 1. Introduction

Tuberculosis (TB) is a major contributor to the global burden of infectious diseases, causing over one million deaths annually. The WHO European Region accounts for 3% of global TB cases, with significant variations in disease burden across countries.[1,2] Controlling the spread of TB remains a top priority for global health efforts. The concept of the “burden of tuberculosis” is used to assess the epidemiological landscape, incorporating data on the incidence and prevalence of the disease.[3,4]

According to the World Health Organization (WHO), 10 million new cases of TB were reported in 2021, resulting in 1.4 million deaths.[5] This factor accounts for 1.86% of the total global Disability-Adjusted Life Years (DALYs) and results in the loss of 2.54% of total life years. Consequently, TB ranks 12th and 11th in terms of global health importance in these respective categories.[6]

The TB epidemic varies globally, affecting all countries and age groups, though 90% of cases occur in adults, with 9% among people with HIV (72% in Africa). Two-thirds of cases are concentrated in India (27%), China (9%), Indonesia (8%), the Philippines (6%), Pakistan (5%), Nigeria (4%), Bangladesh (4%), and South Africa (3%), with 87% of all cases found in these and 22 other WHO high-burden countries. Only 6% of cases are in the WHO European and Americas regions. Developed countries show a decline; in the USA, TB incidence dropped from 9.7 to 2.2 per 100,000 from 1993 to 2020. In South Korea, multidrug-resistant tuberculosis (MDR-TB’s) economic burden varies, with higher DALY rates among middle-aged individuals, while in 2017, TB accounted for 6.5% of the total DALY index, rising to 17% among those over 65.[7,8]

Tuberculosis remains a major public health concern in Central Asia. Collectively, Kazakhstan, Kyrgyzstan, Tajikistan, Turkmenistan, and Uzbekistan report over 34,000 TB cases and 8,000 drug-resistant TB (DR-TB) cases annually.[9]

According to estimates, in 2021, the highest TB incidence rate in the WHO European Region was in Kyrgyzstan (130 per 100,000 population), followed by Tajikistan (88), the Republic of Moldova (85), Kazakhstan (73), and Ukraine (71).[5,10] Armenia, Azerbaijan, and Georgia, the three countries comprising the Southern Caucasus, are among the 18 high-priority countries in eastern Europe and central Asia that account for 85% of TB incidence and over 90% of drug-resistant TB cases in the WHO European Region.[11]

Although there have been substantial decreases in case notifications over the past decade, TB remains a significant public health issue in the South Caucasus. In 2017, Armenia had 812 cases (27.1 per 100,000 people), Azerbaijan had 7,129 (67 per 100,000) and Georgia had 2,927 notified cases (69 per 100,000). The rates in most EU countries is under 10 per 100,000.[12]

TB death rate of Mongolia remained stable at 10.0 cases per 100,000 people over the last 3 years.[13] Mongolia, a lower middle-income country of 3 million people, has the fourth-highest TB incidence in the WHO Western Pacific Region, at 428 cases per 100 000, and among the world’s lowest TB treatment coverage rates, at 31%.[14]

Globally, key risk factors for TB include biological, socioeconomic, and behavioral aspects. HIV co-infection, diabetes, and previous TB treatment significantly increase susceptibility, while poverty, overcrowding, smoking, and alcohol use further elevate the risk.[15] Several studies have examined the influence of risk factors on TB prevalence in different groups. For instance, a study by Daniel E. et al. in Tajikistan identified 59 cases of active pulmonary TB among prisoners, with a point prevalence of 4.5% (95% CI: 3.4–5.7).[16] Similarly, Tilloeva Z. et al. investigated TB among a key population group, reporting that 29.8% of patients had a positive sputum smear, 14.1% were drug-sensitive, and 11.3% had mono-DR/MDR-TB.[17]

The COVID-19 pandemic has had a significant negative impact on efforts to combat TB.[18] Reduced diagnosis and treatment, due to restrictions and overburdened health systems, has led to increased mortality and more severe cases. [19,20] In regions with high TB incidence, the situation has further deteriorated due to the economic repercussions of the pandemic.[21] Additionally, patients co-infected with COVID-19 and TB face an increased risk of death. Logistical challenges, such as rising transport costs, have also complicated the implementation of TB control programs.[20] During the COVID-19 pandemic, there was a sharp increase in air and sea transportation costs, leading to higher import expenses, which may negatively impact TB control programs.[22]

Despite increased costs and a decline in the global incidence of TB from 2000 to 2017, many TB control programs continue to experience funding shortages, particularly in low-and lower-middle-income countries.[5,23] This financial strain is evident in Uzbekistan, where between 2016 and 2020, the annual cost of purchasing medicines for the recommended 6-month treatment course for HPV-TB and the 20-month course for MDR-TB using oral therapy averaged $1.4 million (± $274,000), with an additional $34,000 (± $6,400) per year allocated for the import of medicines.[22] A similar challenge exists in Kazakhstan, where the estimated cost of diagnosing and treating a TB patient for the current episode is $929, while for MDR-TB patients, it rises to $3,125. These financial burdens highlight how the cost of anti-tuberculosis drug regimens can directly impact the number of individuals receiving treatment, potentially limiting access to life-saving therapies.

It is important to note that the situation regarding TB in Central Asia remains a significant problem. Although the absolute number of cases has decreased, the prevalence of TB in the region is still extremely high compared to Europe and America, presenting substantial challenges for both governments and patients. This is partly attributed to the socio-economic conditions of the population and the lack of medical facilities.[24] While some individual studies and global reports on TB prevalence have been conducted, there is no comprehensive systematic review addressing the broader range of TB effects in Central Asia.[24]

Despite a decline in TB incidence in many parts of the world, the burden of TB remains a significant public health challenge in Central Asia, the South Caucasus, and Mongolia. Most of the countries in this study, except Mongolia, were part of the former Soviet Union and share a common healthcare legacy, including centralized TB control programs, diagnostic approaches, and systemic healthcare challenges that persist today. Mongolia, while not a former Soviet republic, had strong political and economic ties with the USSR and adopted a similar healthcare model, making it a relevant case for comparison.

This study focuses on these specific countries because they continue to face high TB prevalence, drug-resistant TB strains, and socio-economic factors that hinder effective TB control.[25] Unlike other post-Soviet states, such as the Baltic countries or Eastern European nations, which have significantly lower TB rates and stronger healthcare infrastructure, the selected regions remain among the most affected. Therefore, the purpose of this systematic review is to analyze the prevalence, risk factors, and healthcare challenges associated with TB in Central Asia, the South Caucasus, and Mongolia, providing a comprehensive regional perspective and country comparison on the ongoing TB epidemic.

## 2. Materials and methods

### 2.1. Study design

An analysis was conducted based on a systematic review of data regarding the prevalence of TB in the Central Asia, Southern Caucasus, and Mongolia. The study was reported in accordance with the PRISMA checklist, which is presented in Table 1 of the supplementary materials.

**Table 1.**
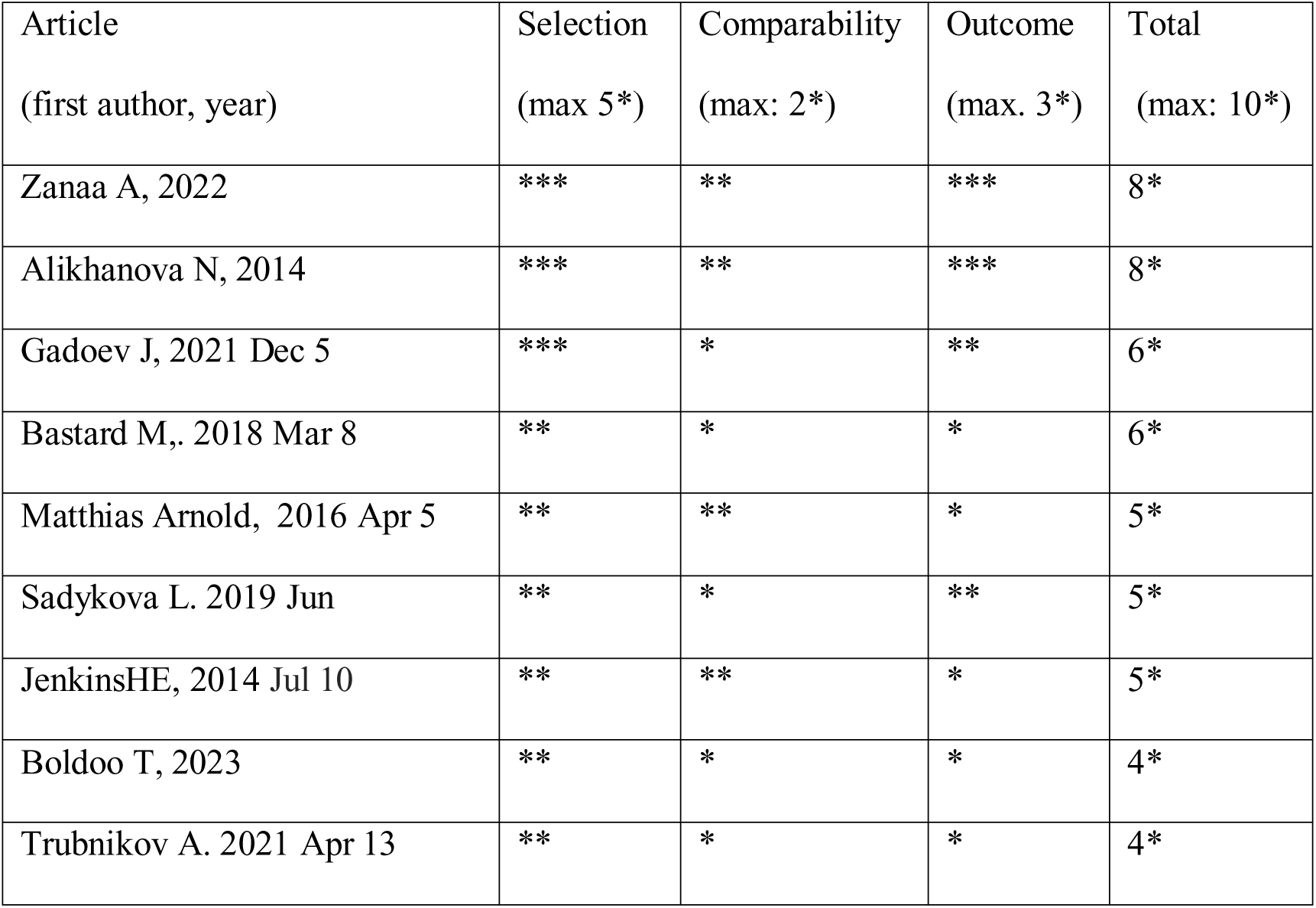
Quality assessment according to the Newcastle-Ottawa scale (NOS), score ≥4, n=9.

### 2.2. Protocol and Registration

The research protocol was registered on the PROSPERO website (https://www.crd.york.ac.uk/PROSPERO/view/CRD420251020764). The registration number is PROSPERO 2025 CRD420251020764.

### 2.3. Eligibility criteria

The results of a study reflecting the prevalence of TB in the Central Asian countries of Kyrgyzstan, Uzbekistan, Tajikistan, Kazakhstan, and Turkmenistan, also in Mongolia and in the Southern Caucasus countries of Armenia, Azerbaijan and Georgia should be presented. The study population should include patients diagnosed with TB. Given the high incidence of TB in the region, studies reporting the prevalence of multidrug-resistant TB (MDR-TB) among TB patients are also acceptable.

The language of the research may be either English or Russian, as international studies are primarily published in English, while national journals in Central Asia and in Southern Caucasus are published in Russian.

The review included full-text scientific articles (excluding conference abstracts) on the prevalence of TB (model, population, or survey), published in peer-reviewed journals or national reports and reviews available from official government sources. All articles published from the beginning of the database search, spanning from 2013 to 2023, were included.

### 2.4. Sources of information

Databases including PubMed and Google Scholar were used for the search. We limited the search to articles published within the last 11 years, from 2013 to 2023.

### 2.5. Search

The following keyword combinations were used in the database search: “prevalence”, “burden”, “economy”, “Tuberculosis”, “Multidrug resistance”, “Pulmonary tuberculosis”, “Mycobacterium tuberculosis”, “Central Asia”, “Southern Caucasus”, “Kazakhstan”, “Tajikistan”, “Kyrgyzstan”, “Uzbekistan”, “Turkmenistan” and “Mongolia”, “Armenia”, “Azerbaijan”, “Georgia”.

Inclusion and exclusion criteria for study selection

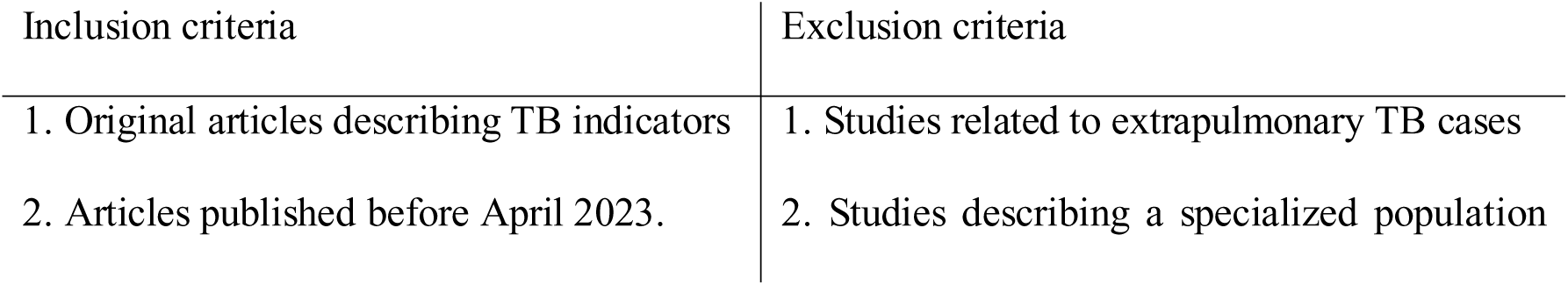
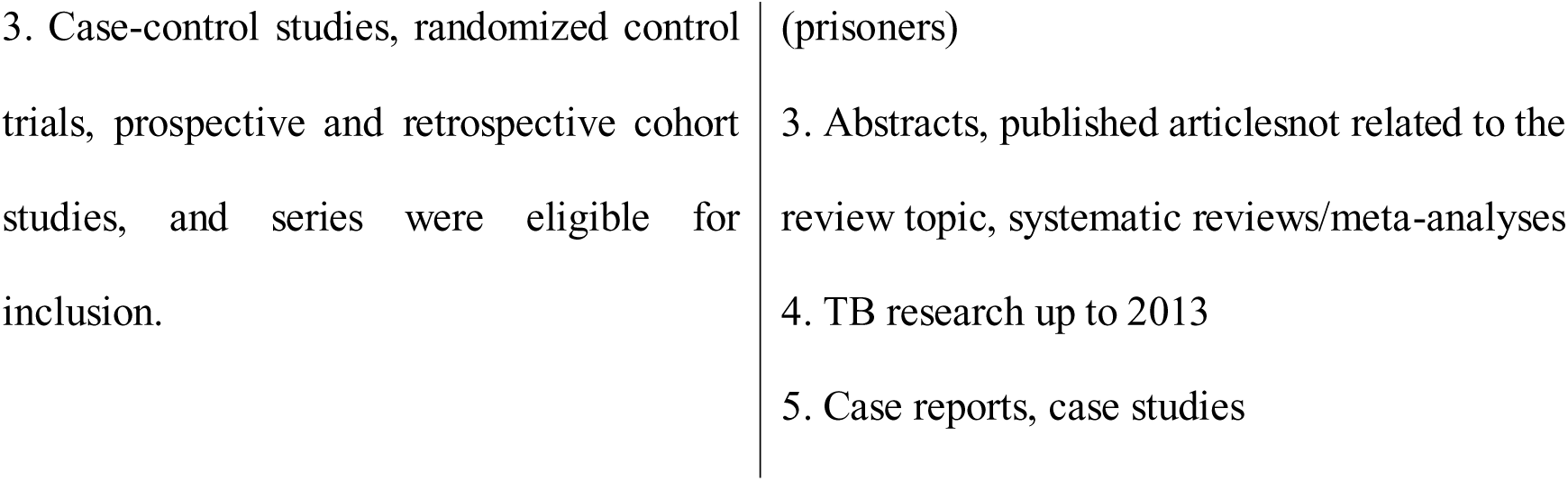

| Inclusion criteria | Exclusion criteria |
| --- | --- |
| 1. Original articles describing TB indicators | 1. Studies related to extrapulmonary TB cases |
| 2. Articles published before April 2023. | 2. Studies describing a specialized population |
| 3. Case-control studies, randomized control trials, prospective and retrospective cohort studies, and series were eligible for inclusion. | (prisoners)<br>3. Abstracts, published articles not related to the review topic, systematic reviews/meta-analyses<br>4. TB research up to 2013<br>5. Case reports, case studies |

### 2.6. Selecting a study

The researchers reviewed the titles and abstracts of all retrieved articles to assess whether they met the predefined criteria for inclusion or exclusion from the study. The full texts of articles that met these criteria, based on the initial review of titles and abstracts, were then carefully examined and reassessed to determine their final inclusion in the study, considering the established criteria. The entire process of selecting articles for the study was documented using the PRISMA flow chart.

### 2.7. Data Elements

The following information was extracted from each eligible article or report: first author details, year of publication, study design, sample size, setting (country), population characteristics (including age and sex), disease characteristics (including type of TB), and disease prevalence. Prevalence data were extracted and reported separately.

### 2.8. Quality assessment

We assessed the quality of the articles using the Newcastle-Ottawa Scale “(NOS)” for cross-sectional studies, as outlined in Table 1. This scale ranges from 0 to 10 points and includes criteria for evaluating selection (up to 5 points), comparability (up to 2 points), and presentation of results (up to 3 points).

#### 2.8.1. Quality of Studies

The quality assessment using the NOS Scale is presented in Table 1. The overall quality was deemed satisfactory, with a mean score of 5.67 (SD >1.46) out of 10 for all studies combined. In the screening section, the mean score was 2.33 (SD ≈ 0.50) out of 5, with only two studies [Zanaa A et al., 2022, and Alikhanova N et al., 2014] achieving the maximum score of 8. In the comparability section, the mean score was 1 (SD ≈ 0.50) out of a possible 2. In the results section, the mean score was 1.89 (SD = 0.78) out of 3. After the quality assessment, 9 articles with a score higher than 4 were included in this review.

### 2.9. Summary measures

The primary outcome measure was TB prevalence. Data were stratified by demographic characteristics, including age, sex, country, and risk factors, as well as healthcare costs, and were presented in a narrative format.

## 3. Results

### 3.1. Study Selection

As a result of the search and selection process (figure 1-PRISMA), 131 publications remained after duplicates were removed and were considered for analysis. Following the application of inclusion criteria, 47 articles were selected, of which 13 were included in the final analysis after applying the exclusion criteria. After assessing the quality of the studies using the NOS scale, 9 articles were selected for detailed analysis.

**Figure 1:**
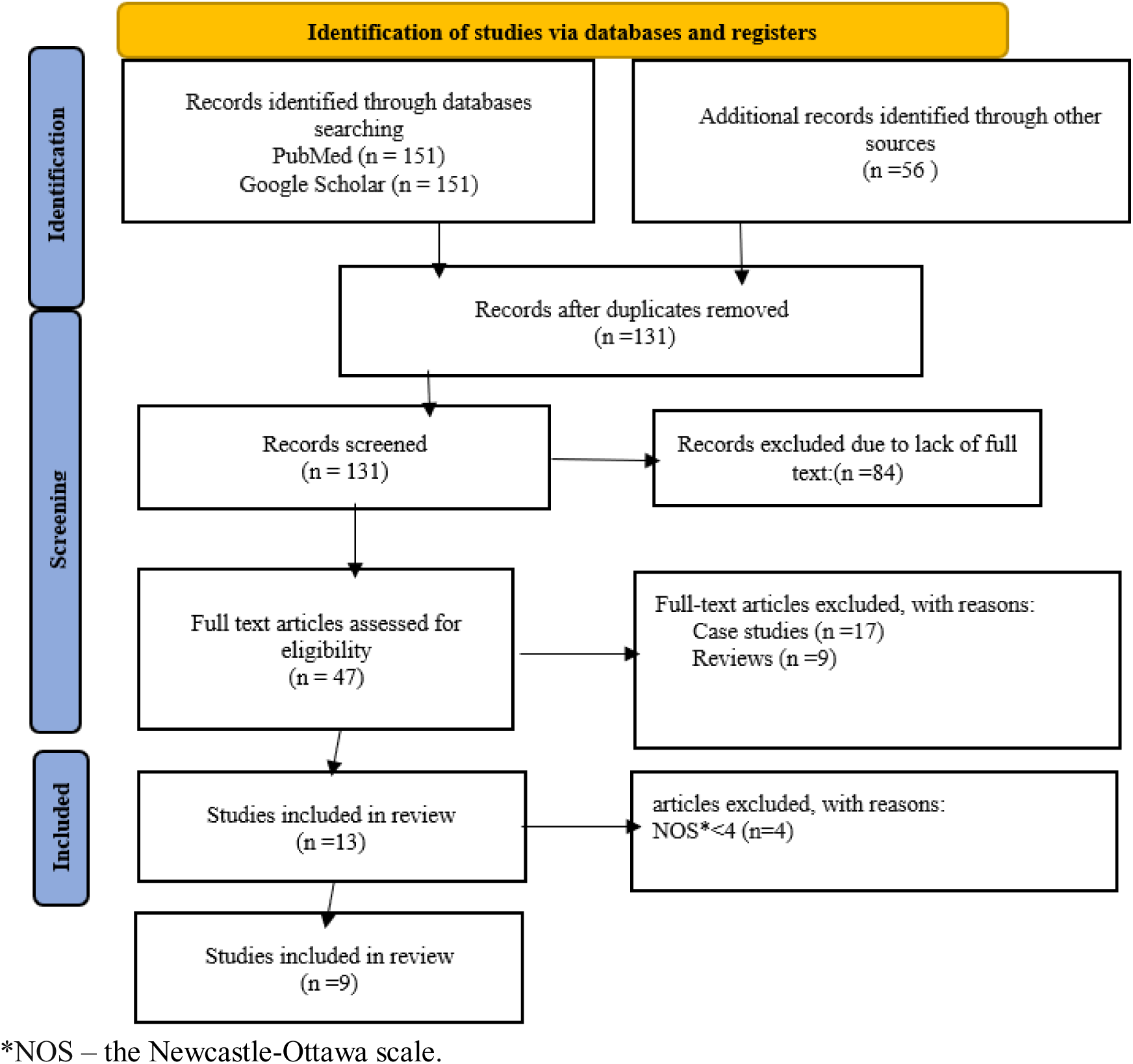
Flowchart of the literature search strategy.

### 3.2. Prevalence of TB in Central Asia

In Central Asia and in Southern Caucasus, studies have been conducted to assess the prevalence of TB, considering demographic characteristics and risk factors. Research on drug-resistant forms of tuberculosis has been carried out in several countries in this region, including Kazakhstan, Uzbekistan, Kyrgyzstan, Armenia, Azerbaijan, and Georgia. Research on drug-resistant forms of TB has been carried out in several countries in this region (Kazakhstan, Uzbekistan, Kyrgyzstan), including some countries of Caucasus (Armenia and Georgia).

The distribution of prevalence study sites is shown in Figure 2.

**Figure 2:**
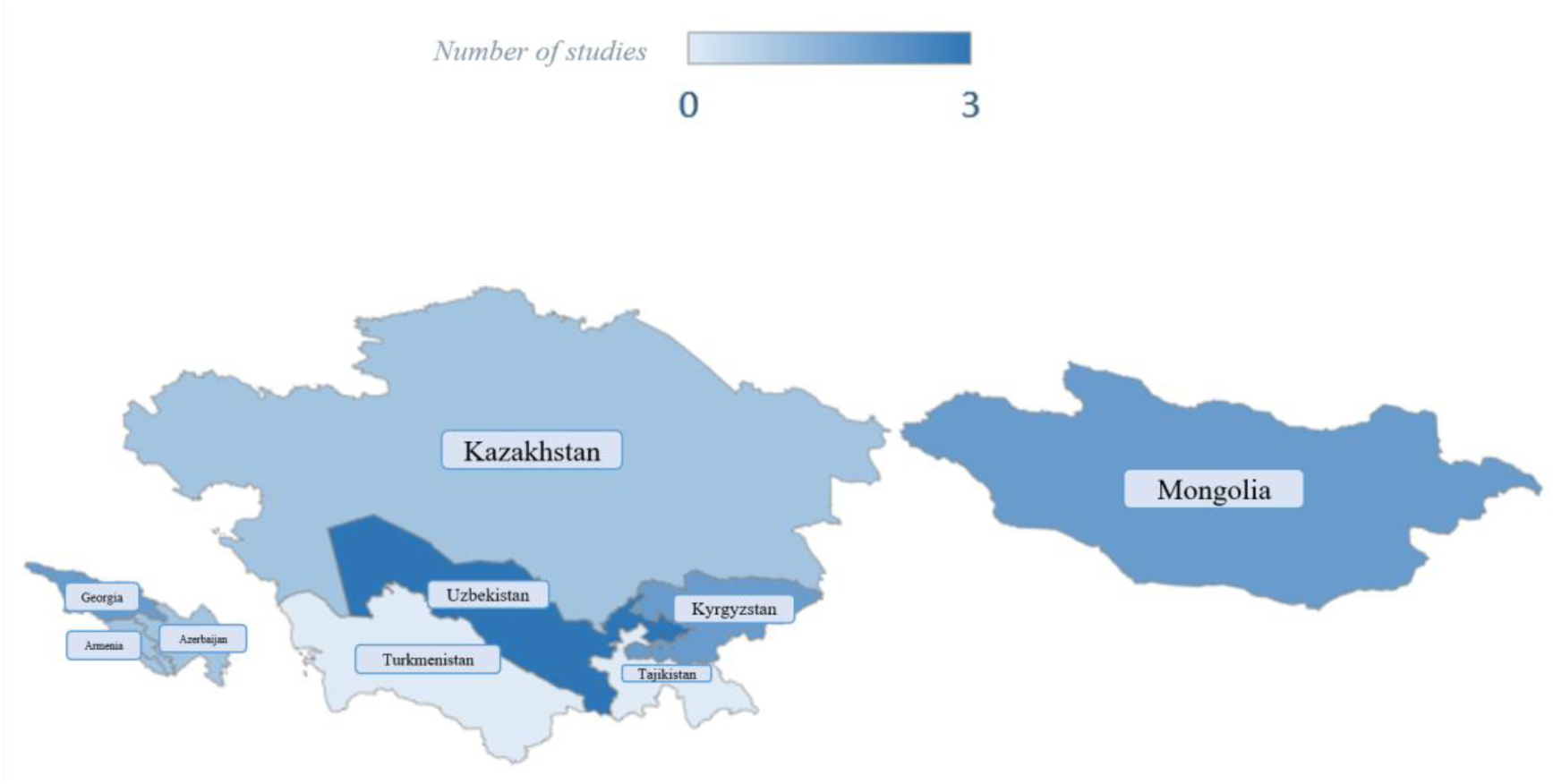
Number of Studies on Tuberculosis Prevalence in Central Asia and the Southern Caucasus. (1) Uzbekistan (3) [[26–29]]; (2) Mongolia, Kyrgyzstan, Georgia (2) [[30]]; (3) Mongolia (among children) [[31]]; (4) Armenia, Kazakhstan, Azerbaijan (1) [[32]]; (5), Tajikistan, Turkmenistan (0).

The main data of all retrieved studies reporting on the prevalence of TB in Central Asia and in Southern Caucasus are presented in Table 2.

**Table 2.**
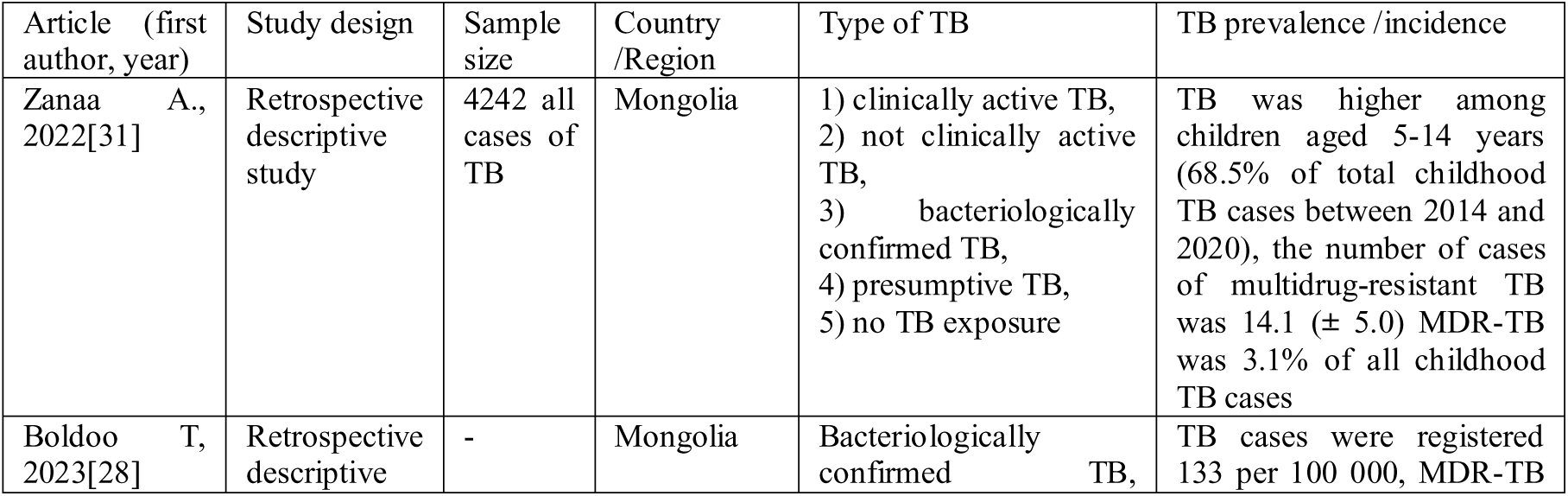
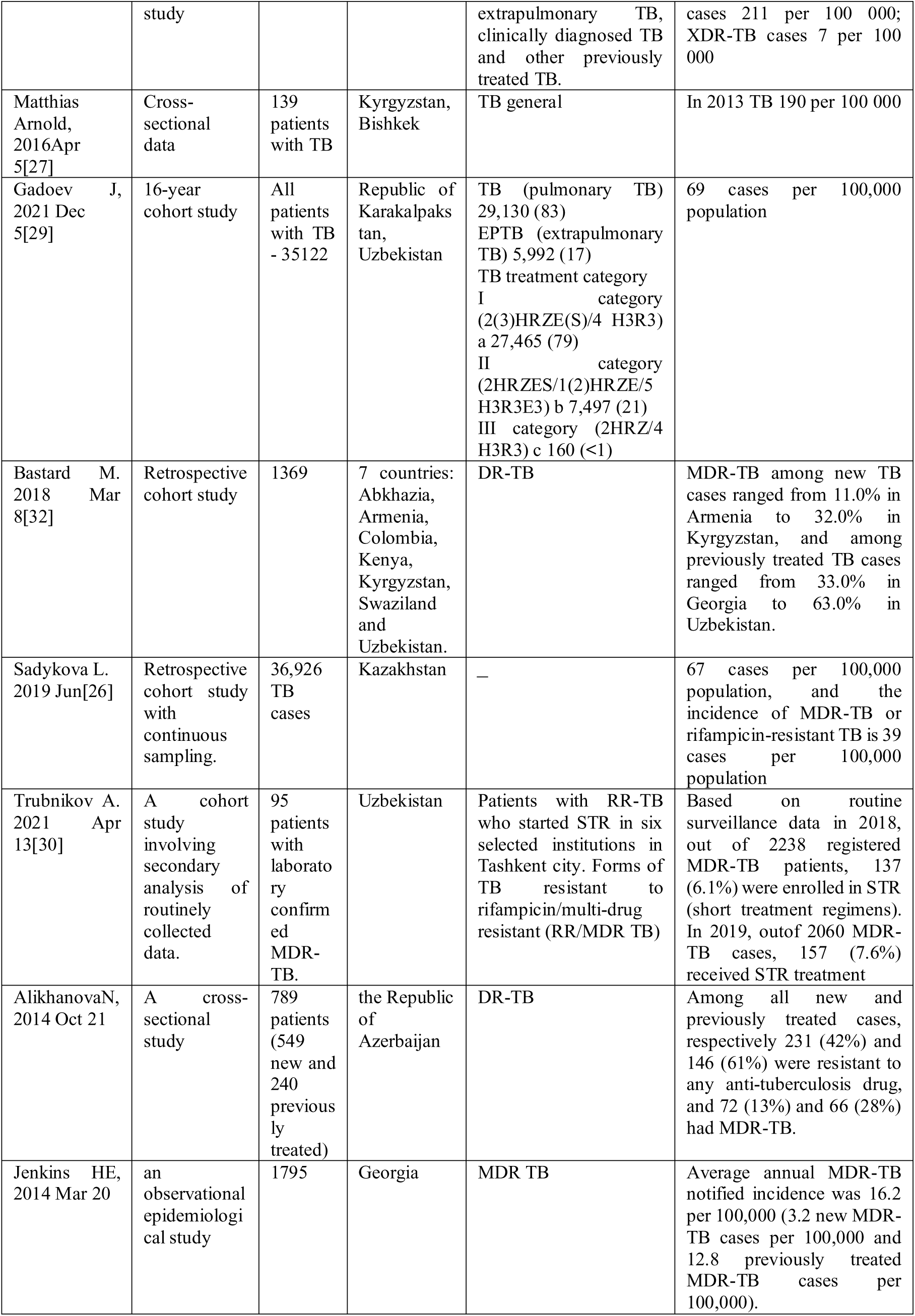
Studies providing data on the prevalence of TB in Central Asia and in Southern Caucasus (n=9).

The study designs demonstrate significant variation, ranging from cross-sectional snapshots, such as Alikhanova N.’s (2014) investigation of 789 TB patients in Azerbaijan, to observational studies like Jenkins HE.’s (2014) analysis of 1,795 TB cases in Georgia. Cross-sectional studies often provide valuable insights into specific TB characteristics, such as drug resistance patterns at a given point, while longitudinal studies are better suited for understanding temporal trends in TB incidence and prevalence.

Sample sizes and geographical coverage also impact the range of findings. Studies with large datasets, such as Gadoev J.’s (2021) examination of 35,122 TB cases in Uzbekistan, offer robust national-level insights, while smaller, targeted investigations like Turbokov’s study on 95 MDR-TB cases in urban Tashkent focus on niche aspects of the TB epidemic. Similarly, Alikhanova’s study in Azerbaijan highlights drug resistance patterns, reporting that among new and previously treated cases, 42% and 61%, respectively, were resistant to anti-TB drugs, and 13–28% had MDR-TB. Meanwhile, Jenkins HE.’s study in Georgia quantified MDR-TB incidence at 16.2 per 100,000 annually, with distinctions between new and previously treated cases.

Regional studies, such as those focused on Karakalpakstan (Uzbekistan) and Bishkek (Kyrgyzstan), provide granular data that capture localized trends and resistance patterns, which may not align with broader national or international analyses. This regional lens can help policymakers identify unique risk factors, but it limits the generalizability of results.

Drug-resistant TB remains a critical focus in many studies, as highlighted by the differences in the prevalence of MDR-TB and DR-TB. While Jenkins HE. emphasizes MDR-TB trends in Georgia, broader studies, such as Bastard M.’s (2018) multi-country analysis, underscore the challenges of managing TB in populations with varying levels of resistance. By focusing solely on resistant strains, such studies may inadvertently exclude broader contextual factors, such as latent TB infections or less severe drug-susceptible cases, which are essential for a complete understanding of the epidemic.

However, the differences in study design, sample size, geographic focus, and types of TB examined across these studies make direct comparisons challenging. These variations highlight the need for a standardized approach in TB research across countries, which would improve comparability and enhance our understanding of regional TB patterns, particularly concerning drug-resistant cases.

### 3.3. Prevalence of TB by sex, age and risk factors

There were seven studies conducted in Central Asia and in Southern Caucasus countries examining the differences in TB prevalence by gender (Table 3). The authors Bastard M., Sadykova L., and Trubnikov A. identified a significant disparity in prevalence between men and women, with averages of 65.3% and 34.7%, respectively.26,32

**Table 3.**
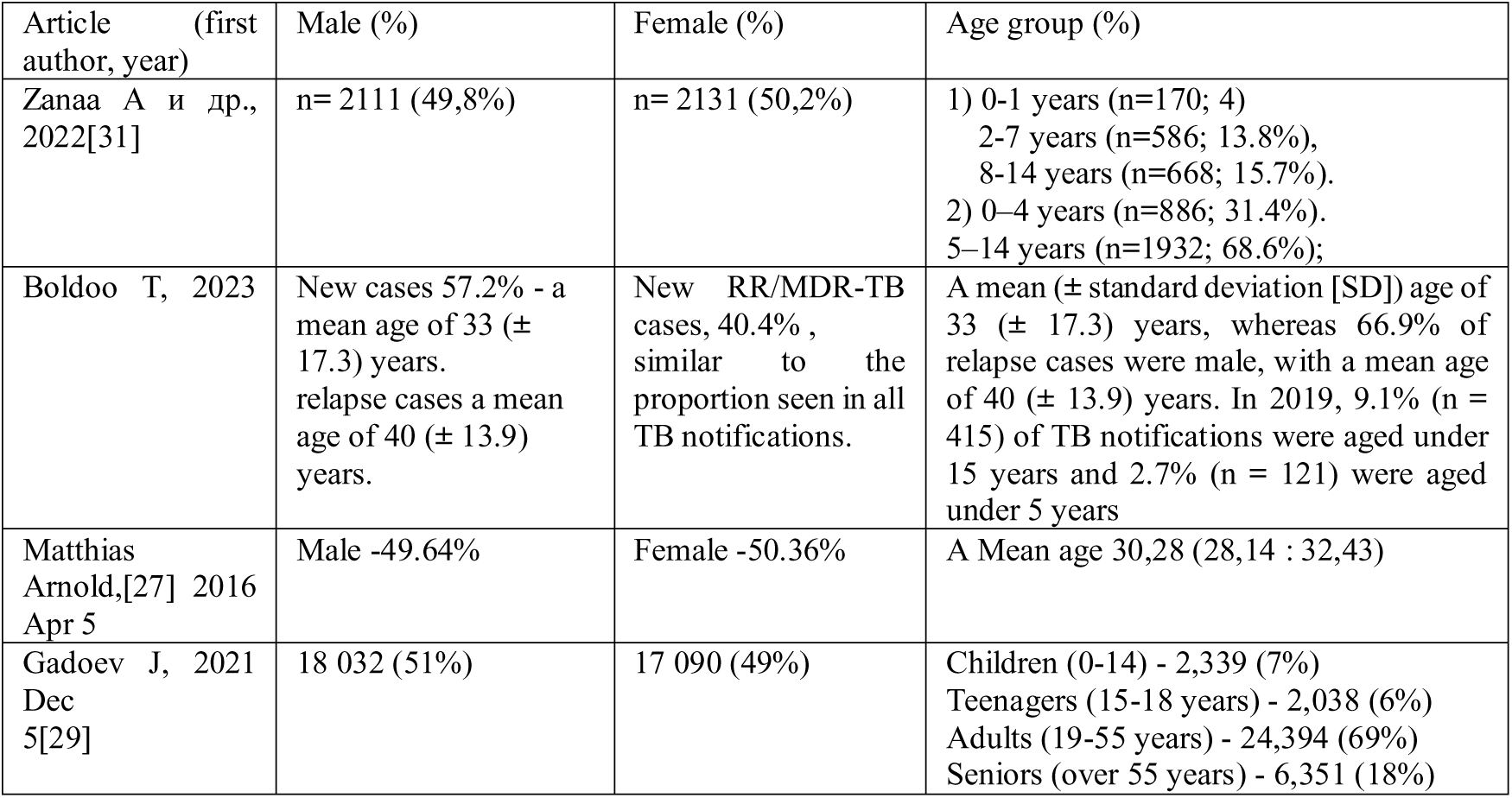
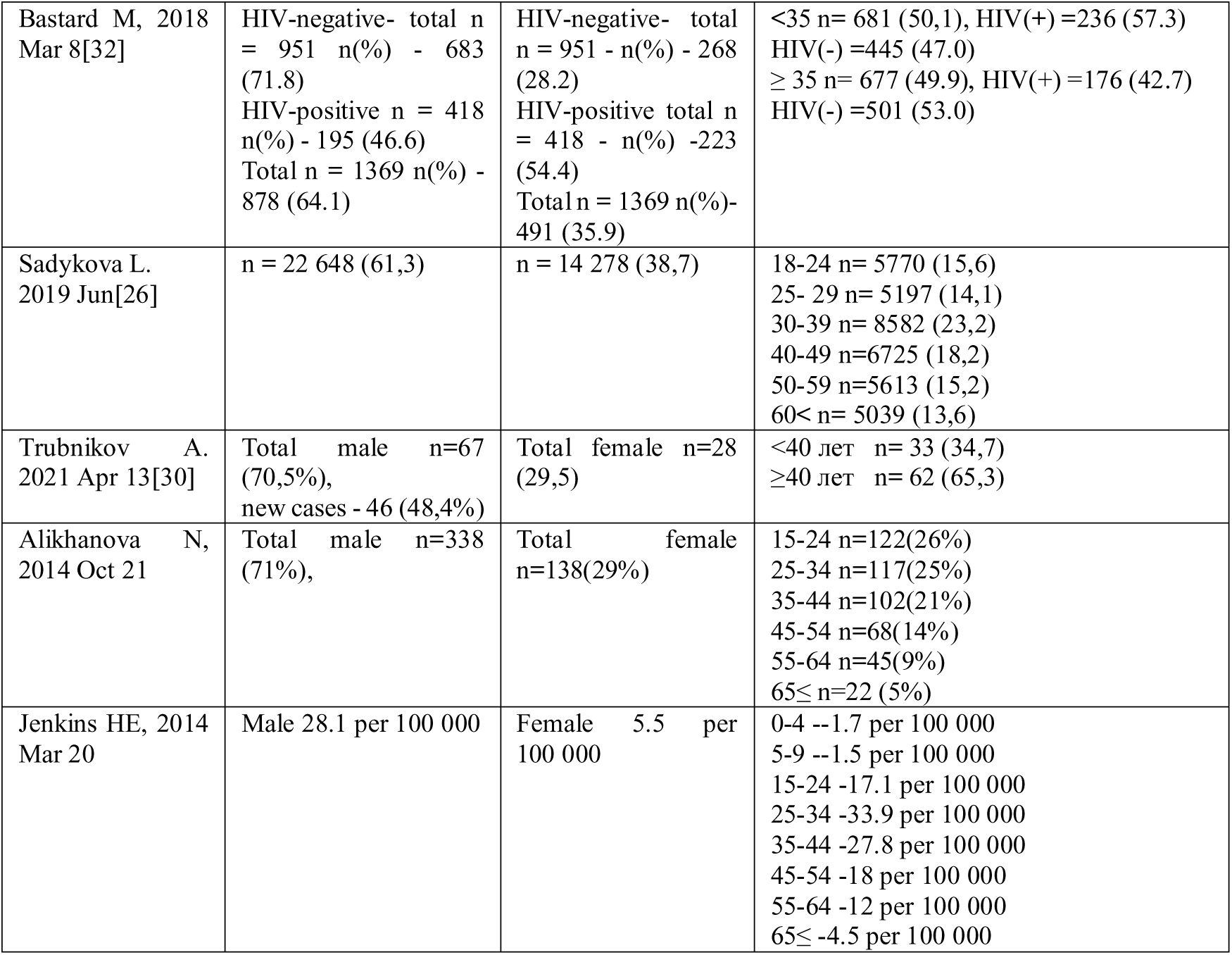
Studies providing data on prevalence of TB by sex and age, n= 9.

The studies indicate gender distribution is generally close to equal, but with slight differences. For example, Zanaa A. (2022) shows a near-equal split (Male 49.8%, Female 50.2%), while Matthias Arnold (2016) shows a similar pattern (Male 49.64%, Female 50.36%). However, Bastard M. (2018) shows a slightly higher male proportion among HIV-negative cases, with males representing 68.3% and females 31.7%, suggesting a potential gender bias in certain subpopulations. Other studies, like Sadykova L. (2019), show almost identical distribution by sex. These small variations highlight the importance of considering biological, social, and behavioral factors that could influence TB prevalence across sexes.

The age groups in these studies differ significantly, complicating direct comparisons of TB prevalence across populations. Each study categorizes age in unique ways, leading to challenges in understanding how TB affects various demographics and making it difficult to aggregate or compare findings accurately.

The inconsistencies in age groupings across studies lead to several comparability issues. For example, Zanaa A. (2022) categorizes early childhood TB cases into narrow groups, such as 0-1 years (170 cases) and 2-4 years (585 cases), which highlights TB prevalence in very young children. In contrast, Sadykova L. (2019) provides detailed intervals for adults, including 18-24 years (5,770 cases), 25-29 years (5,197 cases), and older age groups, which focus more on adult TB prevalence patterns. Studies like BoldooT. (2023) and Matthias Arnold (2016) report only a mean age of TB cases, 33 years (±17.3) and 30.28 years respectively, which doesn’t capture age-specific prevalence patterns as effectively. Differences in age and gender distribution complicate the comparison of TB prevalence across studies. For example, Alikhanova N. (2014) reports 71% of new cases among males and 29% among females, with the majority (72%) occurring in individuals aged 15–44 years. In contrast, Jenkins HE (2014) provides prevalence rates per 100,000 population, showing higher rates in males (28.1) compared to females (5.5), with significant age-specific differences—17.1 per 100,000 in the 15–24 age group and 4.5 per 100,000 in those aged 65 and older.

The results from the reviewed articles indicated that the prevalence of TB is higher in patients over 30 years of age (Table 6). Additionally, studies focusing on the pediatric population reported similar prevalence rates, with an average of n = 1,096 cases among children under 14 years.29,31 These differences hinder comparisons and limit the ability to draw unified conclusions about TB prevalence across life stages. Additionally, variations in age grouping may obscure trends in TB epidemiology, as certain life stages—such as young adulthood (20-39 years, with 8,582 cases) and middle age (40-49 years, with 6,755 cases) in Sadykova’s study—may experience different rates of infection, progression, and treatment outcomes. Studies focusing on relapse cases, like Boldoo T., report an average age of 40 years (±13.9), which is higher than the mean age in new cases, suggesting age-specific patterns in disease recurrence.

To improve comparability, future research should adopt standardized age categories to better capture and compare TB trends across populations, ensuring a clearer understanding of how TB affects various age groups.

Based on the results of the selected articles, groups, and risk factors for TB in Central Asia and in Southern Caucasus countries were identified (Table 4). The authors highlighted socio-economic factors, differences in place of residence, concomitant diseases, HIV status, and contact with TB patients as significant risk factors. High-risk groups included individuals deprived of liberty (prisoners) and those with a positive HIV status. HIV-positive patients accounted for more than half of TB cases, with reported figures of 418 cases (30.5%) by Bastard M. (2018), 490 cases by Sadykova L. (2019), and 88.4% by Trubnikov A. (2021).

**Table 4.**
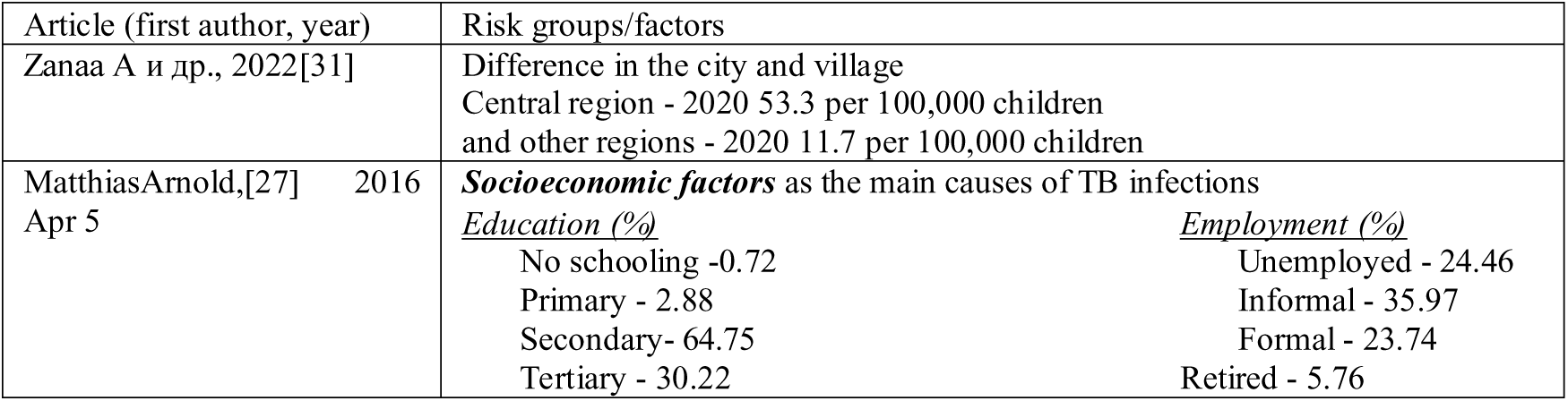
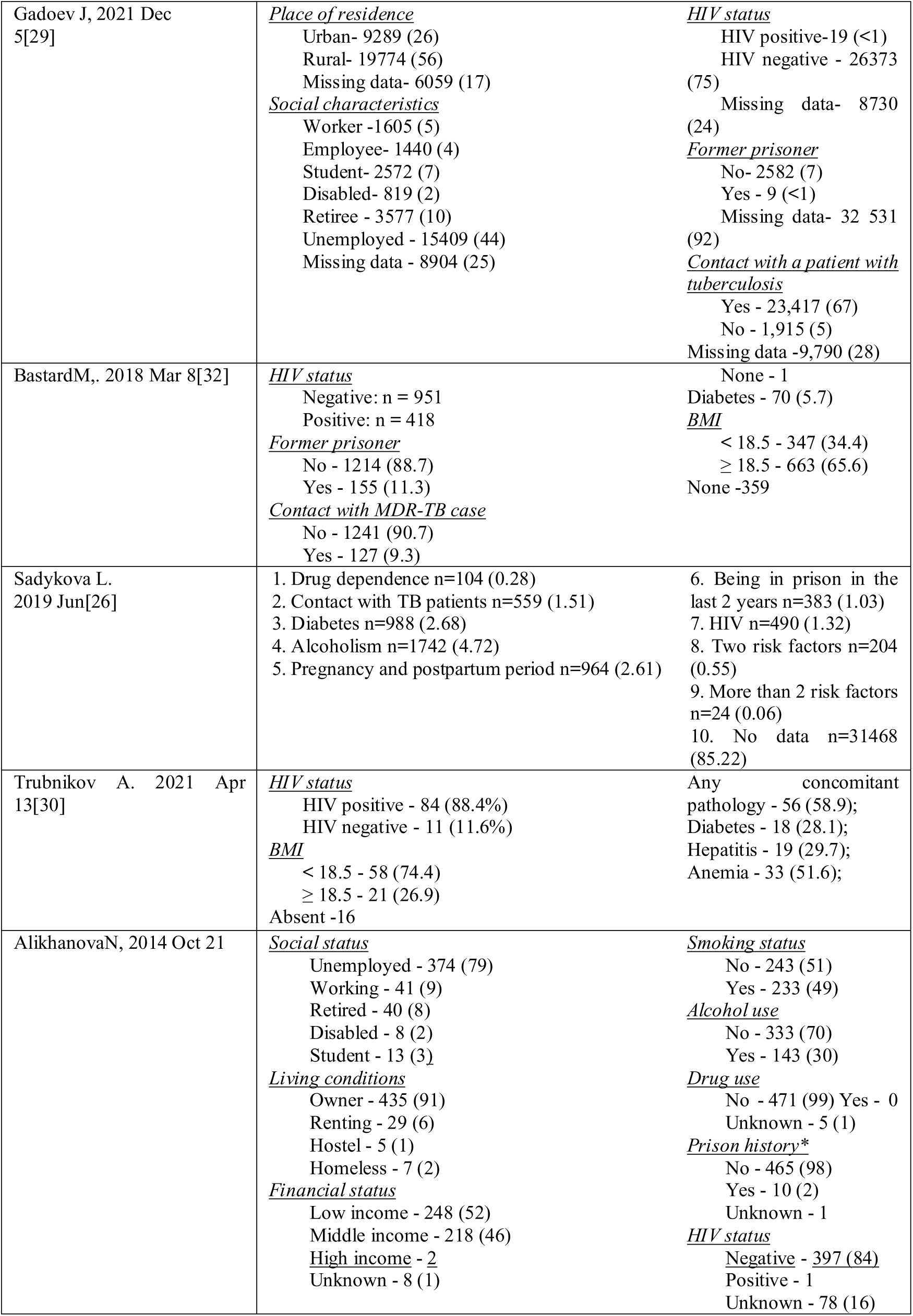
Studies providing data on risk factors for tuberculosis, n= 7.

Additionally, Zanaa A. and Gadoev J. investigated the difference in prevalence among urban and rural residents. Their findings revealed that in Mongolia, the prevalence of TB was higher in urban areas (53.3 per 100,000)31, while in Uzbekistan, a greater number of TB cases were reported in rural areas (19,774 cases, accounting for 56% of the total patient population).29 However, direct comparisons between these countries are not feasible, as the data do not represent the entire population of each country.

### 3.4. Economics of TB

An overview of the only three studies reporting the economic impact of TB is presented in Table 5. Notably, only one study focused on a specific country (Kazakhstan), while the other two were regional studies. The studies generally addressed different aspects and segments of costs, ranging from monthly to episodic expenditures. The cost of treating MDR-TB was significantly higher than that for treating standard TB. Furthermore, we could not find any data on the costs associated with TB treatment in Turkmenistan, Mongolia, and the Southern Caucasus countries.

**Table 5.**
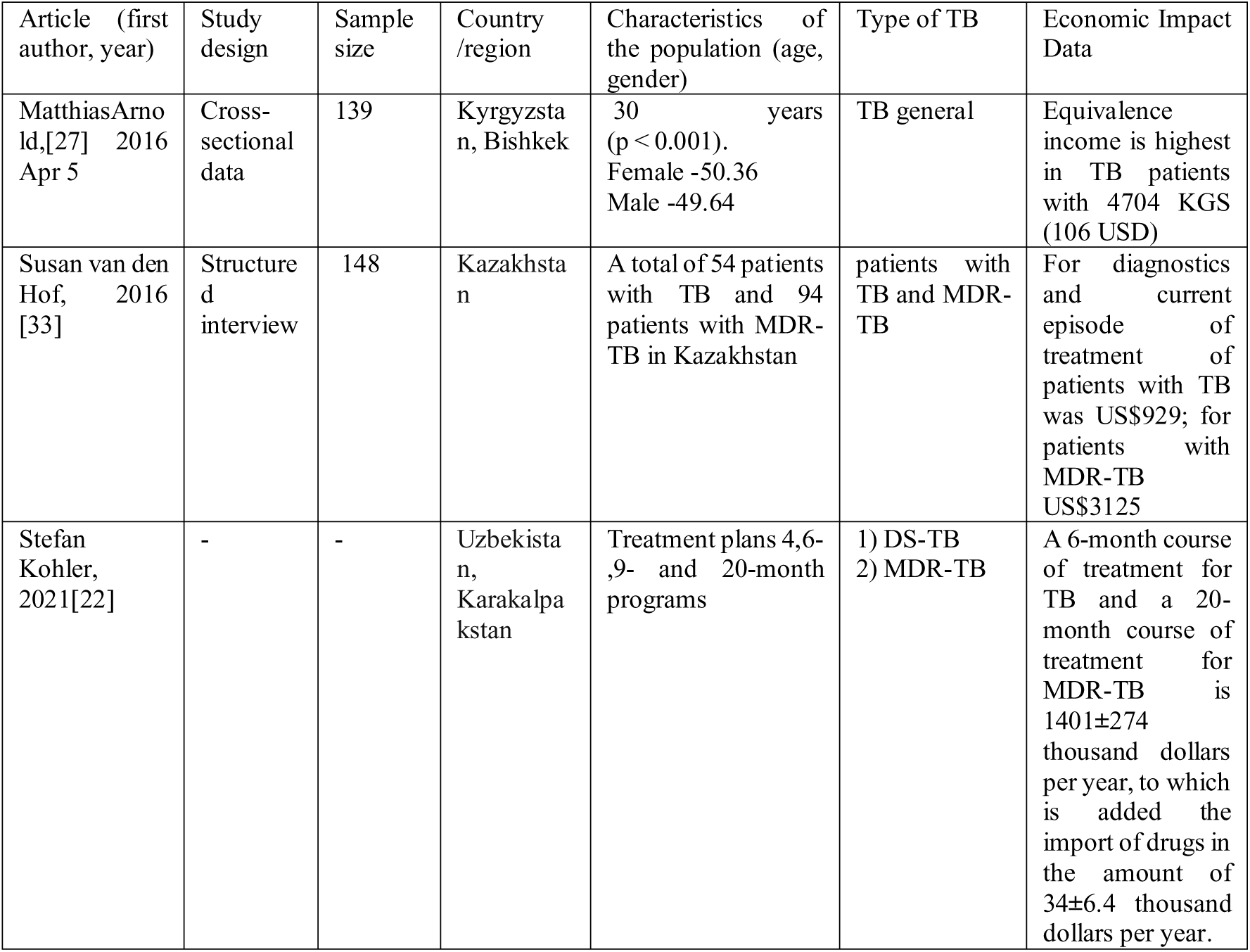
Studies providing data on the cost of tuberculosis treatment in Central Asia, n=3.

## 4. Discussion

We conducted a systematic literature review to identify published data on the prevalence of TB, its associated risk factors and economic challenges in Central Asia, southern Caucasus and Mongolia. Our analysis revealed that 11 studies on the prevalence of TB were conducted in this geographic region. Most of the identified studies (74%) provided data on former Soviet republics such as Kyrgyzstan, Kazakhstan, Uzbekistan, and Tajikistan; however, we did not find data on Turkmenistan, likely because most statistics are considered state secrets and no published data are available.[34]

Our results indicate that the prevalence of TB in Central Asia, southern Caucasus and Mongolia remains high, as reported by the WHO.[5] However, rates vary significantly across countries and territories. For instance, Sakko et al. (2019) report that in Kazakhstan, the incidence of TB decreased from 227 to 15.2 cases per 100,000 people, while all-cause mortality among TB patients rose from 8.4 to 15.2 per 100,000 between 2014 and 2019.[35] TB incidence in Mongolia showed an increasing trend from 41.65 to 55.63 per 100,000 between 2016 and 2018.[36] However, in our review, TB incidence rates in Mongolia declined from 2015 to 2018, but increased again in 2019 to 133 per 100,000.[28] However, the heterogeneity of the data presented in the articles poses challenges in drawing comprehensive conclusions at the national level. The prevalence of MDR-TB ranged from 19% to 26% among newly treated TB patients and from 60% to 70% among previously treated TB patients. Based on the results, it can be concluded that there is a need for evidence focused on identifying and outlining vulnerable population groups.

Analysis of notification rates from 2000 to 2017 reveals diverse trends in drug-resistant TB among these countries. While Armenia and Georgia showed stable notification rates for drug-resistant TB among new cases in recent years, Azerbaijan experienced an increase.[11] In 2017, Armenia had 812 cases (27.1 per 100,000 people), Azerbaijan had 7,129 (67 per 100,000) and Georgia had 2,927 notified cases (69 per 100,000). The rates in most EU countries is under 10 per 100,000.[12]

Factors that were more likely to be associated with TB infection included age > 30 years, male sex, urban residence, and previous TB infection.[37]

In Central Asia, TB incidence is higher among middle-aged individuals (ages 20-55), whereas in other countries, such as Brazil, it is more prevalent among younger populations. The odds of acquiring TB increased significantly with age in young females (ages 5–14 years), while in young adult males (ages 15–39 years), the increase in odds was marginally significant.[38] A study conducted in Myanmar found that the highest positive results by age were observed among individuals aged 16–20 years.[39] Similarly, in Georgia, the median age at death for individuals who died following TB treatment was 64.0 years.[40]

Sex and age were shown to be important risk factors for TB prevalence in the literature we revised. These factors are also often discussed in other studies across the world. For instance, it was shown that gender differences in prevalence among TB patients have been showed that the rates were higher in the male population compared to females globally, averaging 9.39 per 100,000 and 7.95 per 100,000, respectively.[41] In most age groups, males had higher disability-adjusted life year (DALY) rates than females, with the burden of MDR-TB increasing with age.[41] Gender may influence the development and progression of TB due to differences in social roles, risk behaviors, and activities. Alcohol and tobacco dependence tend to be higher in men, which exacerbates the initial clinical presentation at the time of TB diagnosis.[42]

TB has been found to be more prevalent in areas with high population density.[43] Numerous studies indicate that population density is a significant factor influencing the prevalence of childhood TB.[31,43,44] In Uzbekistan, TB is most commonly reported in rural areas among adults.[29] Globally, the prevalence of TB is considerably higher in urban areas than in rural areas.[45] However, in some countries, these statistics are reversed, with individuals residing in rural areas at much higher risk of contracting TB disease. Similarly, in other developing countries where large portions of the population are rurally located, TB incidence in rural areas is greater than or equal to that in large urban locales. So previous studies in the South Africa revealed that rural populations are significantly more likely to experience higher levels of transmission compared to their urban counterparts. Urban residents often live closer to healthcare facilities and have easier access to TB treatment and care, which can reduce the duration of the disease. Additionally, rural populations may have fewer venues for social gatherings (e.g., churches, alcohol-related establishments), leading to a higher concentration of individuals in limited locations, which may contribute to increased transmission compared to urban settings.[46] The prevalence of several co-existing illnesses or risk factors was higher in the TB cohort than in the overall Georgia population in 2014 (our study midpoint), including HIV (10% vs. 0.5%), homelessness (10% vs. 0.12%), excess alcohol use (15% vs. 5.3%), and diabetes mellitus (12% vs. 11%).[40]

During the Covid-19 pandemic, globally there has seen a significant decline in the detection of TB cases. According to experts, the decrease in the number of new cases is not due to a decrease in the incidence of the disease, but rather to incomplete detection of cases. As a result, there is every reason to believe that the number of cases may increase in the coming years.[47] As the coronavirus pandemic impacted people’s lifestyles in 2020, the number of confirmed TB cases dropped significantly before rising rapidly again in 2021.[48]

Our systematic review found that little has been studied in Central Asia about the impact of COVID-19 on TB incidence or mortality (suppl.file table 3). But in Mongolia, Turkmenistan and in the Southern Caucasus countries research on the impact of the pandemic on TB could not be found.

However, Gabdullina M. and colleagues found that during the COVID-19 pandemic in Kazakhstan, there was a significant change in TB rates compared to previous years. The incidence rate was 39%, compared with 31% in previous periods, the proportion of individuals aged 60 and older diagnosed with TB increased significantly from 16% before the COVID-19 pandemic to 22% during the pandemic especially. The pandemic was accompanied by an increase in adverse outcomes from 11% to 20%, as well as an increase in mortality from 6% to 9%.[49] Tajikistan has seen a 28 percent decrease in the number of registered TB cases in 2020 compared to 2019, leading to a decrease in TB treatment coverage.[50]

Globally, there was a 3% increase in drug-resistant tuberculosis (DR-TB) cases between 2020 and 2021. There were 450,000 new cases of rifampicin-resistant tuberculosis (RR-TB) in 2021.[51]

SachinSilva and co-authors examined the economic impact of TB mortality in 120 countries. From 2020 to 2050, with the current 2% annual decline in TB mortality, there would be an estimated 31.8 million TB deaths (95% uncertainty interval: 25.2 to 39.5 million), resulting in economic losses of US$17.5 trillion (US$14.9 to 20.4 trillion). However, if the SDG target for TB mortality is met by 2030, an estimated 23.8 million TB deaths (US$18.9 to 29.5 million) and economic losses of US$13.1 trillion (US$11.2 to 15.3 trillion) could be avoided.[52]

However, we could not find sufficient evidence on the economic impact of TB prevalence in the countries of interest. There is a lack of in-depth data on the costs associated with TB treatment, including direct medical expenses, indirect costs, and the broader socioeconomic consequences. Addressing these gaps through further research is crucial for developing effective policies and resource allocation strategies.

## 5. Strengths and Limitations

The strength of this study is that it is the first systematic review of TB prevalence in Central Asia, Southern Caucasus and Mongolia.

Аn extensive literature review was conducted by two independent reviewers in accordance with PRISMA guidelines. Furthermore, the protocol for this study was registered in advance with PROSPERO, and we included articles from peer-reviewed journals in both English and Russian. The Newcastle-Ottawa Scale was utilized for quality assessment.

Our study identified several limitations: the lack of data on the prevalence of TB in Turkmenistan significantly reduces the objectivity and representativeness of the analysis results. Additionally, the diversity of methodologies and study designs employed across different countries poses challenges for generalizing the findings. Despite the broad scope of the literature search, the total number of available studies may be insufficient to draw comprehensive conclusions for the region as a whole.

## 6. Conclusions

In conclusion, TB incidence in Central Asia, Southern Caucasus and Mongolia varies considerably across countries, with some regions experiencing significantly higher rates than others. The burden of TB is more pronounced among men than women, and the disease predominantly affects individuals of working age, with fewer cases reported in children and a moderate prevalence among the elderly.

Despite a decrease in the absolute number of cases, TB prevalence in the region remains exceptionally high compared to Europe and America, continuing to pose a major concern. This situation necessitates substantial financial expenditures from both governments and patients, largely due to the socio-economic challenges of the population.

Key risk factors for TB transmission include HIV-positive status, comorbidities such as diabetes, and undernutrition. Social determinants like unemployment, imprisonment, and close contact with TB patients also significantly contribute to the spread of the disease.

The economic burden of TB, particularly multidrug-resistant TB (MDR-TB), is substantial, with treatment costs straining healthcare systems across the region. The financial impact varies by country, with higher costs associated with managing drug-resistant strains of TB. These trends underscore the urgent need for more effective prevention, early detection, and management strategies to reduce both the health and economic impacts of TB in Central Asia, Southern Caucasus and Mongolia.

## 7. Recommendations for future research and policy

Despite a decrease in cases, TB prevalence in the studies countries remains high compared to Europe and America, requiring significant financial resources due to the population’s socio-economic challenges. Further research is recommended to expand studies on TB in Central Asia, South Caucasus and Mongolia, particularly by including Turkmenistan, to gain a more comprehensive understanding of TB dynamics in the region. Additionally, more in-depth investigations are needed to assess the impact of factors such as poverty, healthcare access, and social conditions on TB prevalence.

To enhance TB control efforts, it is crucial to establish reliable registers that provide accurate data on TB prevalence, improve data comparability between countries, and better identify vulnerable groups. These measures will contribute to the successful recognition and treatment of TB. Furthermore, ongoing research is essential to examine the long-term effects of the COVID-19 pandemic on TB diagnosis, treatment, and outcomes, ensuring the development of effective interventions to reduce TB mortality in the post-pandemic period.

## Data Availability

All data produced in the present study are available upon reasonable request to the authors Malika Idayat

## Acknowledgments

We would like to thank our colleague Gauhar Dunenova for her valuable comments on previous versions of the article. Their input contributed substantially to the improvement of the paper.

## Availability of data and materials

The datasets used and analysed during the current study are available from the corresponding author on reasonable reques.

## Author Contributions

Conceptualization, E.VdL., N.G.; methodology, E.VdL., N.G.; software, O.A., K.A., M.I.; validation – M.I., N.G.; formal analysis, M.I., N.K.; investigation, N.G., E.VdL.; resources, M.I., O.A., K.A.; visualization – M.I.; data curation – M.I., N.K.; writing— original draft preparation – M.I., N.G.; writing—review, editing M.I., N.G., E.VdL.; supervision, N.G., E.VdL.; project administration, N.G.; funding acquisition E.VdL.

All authors have read and agreed to the published version of the manuscript.

## List of abbreviations

COVID-19: COronaVIrus Disease 2019
TB: TuBerculosis
HIV: Human Immunodeficiency Virus
WHO: World Health Organization
DALYs: Disability-Adjusted Life Years
USA: United States of America
DR-TB: Drug-Resistant Tuberculosis
MDR-TB: Multidrug-Resistant Tuberculosis
NOS: Newcastle-Ottawa Scale
SD: Standard Deviation
EU countries: European Union countries

## References

1. Tuberculosis. https://www.who.int/news-room/fact-sheets/detail/tuberculosis.

2. Tuberculosis surveillance and monitoring in Europe, 2019. https://www.ecdc.europa.eu/en/publications-data/tuberculosis-surveillance-and-monitoring-europe-2019.

3. Vasilyeva, I. A., Belilovsky, E. M., Borisov, S. E. & Sterlikov, S. А. INCIDENCE, MORTALITY AND PREVALENCE AS INDICATORS OF TUBERCULOSIS BURDEN IN WHO REGIONS, COUN TRIES OF THE WORLD AND THE RUSSIAN FEDERATION. PART 2. TUBERCULOSIS MORTALITY. Tuberculosis and lung diseases 95, 8–16 (2017).

4. Ledesma, J. R. et al. Global, regional, and national sex differences in the global burden of tuberculosis by HIV status, 1990–2019: results from the Global Burden of Disease Study 2019. Lancet Infect Dis 22, 222–241 (2022).

5. GLOBAL TUBERCULOSIS REPORT 2021. (2021).

6. Jarde, A. et al. Prevalence and risks of tuberculosis multimorbidity in low-income and middle-income countries: a meta-review. BMJ Open 12, e060906 (2022).

7. World Health Organization. Global Tuberculosis Report 2018.

8. Skrahin, A. http://qims.amegroups.com/article/view/11254/12011. Quant Imaging Med Surg 6, 338–341 (2016).

9. WHO-USAID project to end drug-resistant tuberculosis in Central Asia concludes. https://www.who.int/europe/news-room/20-10-2023-who-usaid-project-to-end-drug-resistant-tuberculosis-in-central-asia-concludes.

10. Tuberculosis surveillance and monitoring in Europe. WHO; ECDC https://www.ecdc.europa.eu/sites/default/files/documents/tuberculosis-surveillance-monitoring-2023.pdf (2023).

11. Dadu, A., Hovhannesyan, A., Ahmedov, S., van der Werf, M. J. & Dara, M. Drug-resistant tuberculosis in eastern Europe and central Asia: a time-series analysis of routine surveillance data. Lancet Infect Dis 20, 250–258 (2020).

12. Protect Migrants from TB.

13. Mongolia Tuberculosis death rate, 1960-2024- knoema.com. https://knoema.com/atlas/Mongolia/topics/Health/Risk-factors/Tuberculosis-death-rate.

14. Saranjav, A. et al. Assessing the quality of tuberculosis care using routine surveillance data: a process evaluation employing the Zero TB Indicator Framework in Mongolia. BMJ Open 12, e061229 (2022).

15. Millet, J. P. et al. Factors that influence current tuberculosis epidemiology. European Spine Journal 22, 539 (2012).

16. Winetsky, D. E. et al. Prevalence, Risk Factors and Social Context of Active Pulmonary Tuberculosis among Prison Inmates in Tajikistan. PLoS One 9, 86046 (2014).

17. Tilloeva, Z. et al. Tuberculosis in key populations in Tajikistan - a snapshot in 2017. J Infect Dev Ctries 14, 94S–100S (2020).

18. World Health Organization. Global tuberculosis report 2019. 283.

19. Impact of the COVID-19 pandemic on TB detection and mortality in 2020. https://www.who.int/publications/m/item/impact-of-the-covid-19-pandemic-on-tb-detection-and-mortality-in-2020.

20. Davies, M.-A. HIV and risk of COVID-19 death: a population cohort study from the Western Cape Province, South Africa. medRxiv (2020) doi:10.1101/2020.07.02.20145185.

21. Glaziou, P. Predicted impact of the COVID-19 pandemic on global tuberculosis deaths in 2020. medRxiv 2020.04.28.20079582 (2020) doi:10.1101/2020.04.28.20079582.

22. Kohler, S., Sitali, N., Achar, J. & Paul, N. Programme costs of longer and shorter tuberculosis drug regimens and drug import: a modelling study for Karakalpakstan, Uzbekistan. ERJ Open Res 8, 00622–02021 (2022).

23. Micah, A. E. et al. Health sector spending and spending on HIV/AIDS, tuberculosis, and malaria, and development assistance for health: progress towards Sustainable Development Goal 3. Lancet 396, 693 (2020).

24. Tabyshova, A. et al. Prevalence and Economic Burden of Respiratory Diseases in Central Asia and Russia: A Systematic Review. Int J Environ Res Public Health 17, 7483 (2020).

25. Cox, H. S. et al. Multidrug-resistant Tuberculosis in Central Asia. Emerg Infect Dis 10, 865 (2004).

26. Sadykova, L. et al. A retrospective analysis of treatment outcomes of drug-susceptible TB in Kazakhstan, 2013-2016. Medicine 98, e16071 (2019).

27. Arnold, M. et al. Coping with the economic burden of Diabetes, TB and co-prevalence: evidence from Bishkek, Kyrgyzstan. BMC Health Serv Res 16, (2016).

28. Boldoo, T. et al. Epidemiology of tuberculosis in Mongolia: analysis of surveillance data, 2015-2019. Western Pac Surveill Response J 14, (2023).

29. Gadoev, J. et al. Recurrent tuberculosis and associated factors: A five - year countrywide study in Uzbekistan. PLoS One 12, (2017).

30. Trubnikov, A. et al. Effectiveness and Safety of a Shorter Treatment Regimen in a Setting with a High Burden of Multidrug-Resistant Tuberculosis. Int J Environ Res Public Health 18, (2021).

31. Zanaa, A. et al. Childhood Tuberculosis in Mongolia: Trends and Estimates, 2010-2030. Tohoku J Exp Med 257, 193–203 (2022).

32. Bastard, M. et al. Outcomes of HIV-infected versus HIV-non-infected patients treated for drug-resistance tuberculosis: Multicenter cohort study. PLoS One 13, (2018).

33. van den Hof, S. et al. The socioeconomic impact of multidrug resistant tuberculosis on patients: results from Ethiopia, Indonesia and Kazakhstan. BMC Infect Dis 16, (2016).

34. Rechel, B. & McKee, M. The effects of dictatorship on health: the case of Turkmenistan. BMC Med 5, 21 (2007).

35. Sakko, Y. et al. Epidemiology of tuberculosis in Kazakhstan: data from the Unified National Electronic Healthcare System 2014–2019. BMJ Open 13, e074208 (2023).

36. Jing, C. et al. Disease burden of tuberculosis and post-tuberculosis in Inner Mongolia, China, 2016-2018 - based on the disease burden of post-TB caused by COPD. BMC Infect Dis 23, (2023).

37. Selvaraju, S. et al. Prevalence and factors associated with tuberculosis infection in India. J Infect Public Health 16, 2058–2065 (2023).

38. Fernandes, P. et al. Sex and age differences in *Mycobacterium tuberculosis* infection in Brazil. Epidemiol Infect 146, 1503–1510 (2018).

39. Seifert, M. et al. Age and sex distribution of Mycobacterium tuberculosis infection and rifampicin resistance in Myanmar as detected by Xpert MTB/RIF. BMC Infect Dis 21, 781 (2021).

40. Gorvetzian, S., Pacheco, A. G., Anderson, E., Ray, S. M. & Schechter, M. C. Mortality Rates after Tuberculosis Treatment, Georgia, USA, 2008–2019 - Volume 30, Number 11—November 2024 - Emerging Infectious Diseases journal - CDC. Emerg Infect Dis 30, 2261–2270 (2024).

41. Lv, H. et al. Global prevalence and burden of multidrug-resistant tuberculosis from 1990 to 2019. BMC Infect Dis 24, 243 (2024).

42. Min, J. et al. Differential effects of sex on tuberculosis location and severity across the lifespan. Sci Rep 13, 6023 (2023).

43. Wang, X. et al. Spatiotemporal epidemiology of, and factors associated with, the tuberculosis prevalence in northern China, 2010–2014. BMC Infect Dis 19, (2019).

44. Wulan Sumekar Rengganis Wardani, D. & Prasetyo Wahono, E. Spatial Analysis of Childhood Tuberculosis and Social Determinants in Bandar Lampung. doi:10.1051/e3sconf/202020212006.

45. RURAL POPULATIONS.

46. Smith, J. P. et al. Characterizing tuberculosis transmission dynamics in high-burden urban and rural settings. Sci Rep 12, 6780 (2022).

47. Н.А.Алтымышева. Влияние COVID-19 на туберкулез. Здравоохранение Кыргызстана научно-практический журнал 2023, № 2 44–48 (2023) doi:10.51350/zdravkg2023.2.6.7.44.48.

48. Ryckman, T. et al. Ending tuberculosis in a post-COVID-19 world: a person-centred, equity-oriented approach. Lancet Infect Dis 23, e59–e66 (2023).

43. Gabdullina, M. et al. COVID-19 pandemic and other factors associated with unfavorable tuberculosis treatment outcomes—Almaty, Kazakhstan, 2018–2021. Front Public Health 11, 1247661 (2023).

50. Usaid. TAJIKISTAN TB RECOVERY PLAN TO MITIGATE THE IMPACT OF COVID-19.

51. Рост смертности и заболеваемости туберкулезом во время пандемии COVID-19. https://www.who.int/news/item/27-10-2022-tuberculosis-deaths-and-disease-increase-during-the-covid-19-pandemic.

52. Silva, S., Arinaminpathy, N., Atun, R., Goosby, E. & Reid, M. J. Economic Impact of TB Mortality in 120 Countries and What It Will Cost If We Don’t Achieve the End TB Targets: A Full-Income Analysis. SSRN Electronic Journal (2020) doi:10.2139/SSRN.3725628.

